# Limited immune responses after three months of BNT162b2 vaccine in SARS-CoV-2 uninfected elders living in long-term care facilities

**DOI:** 10.1101/2021.11.18.21266502

**Authors:** Macedonia Trigueros, Edwards Pradenas, Dolors Palacín, Carlos Ávila-Nieto, Benjamin Trinité, Josep Maria Bonet-Simó, Mar Isnard, Nemesio Moreno, Silvia Marfil, Carla Rovirosa, Teresa Puig, Eulàlia Grau, Anna Chamorro, Ana Martinez, Ruth Toledo, Marta Font, Jordi Ara, Jorge Carrillo, Lourdes Mateu, Julià Blanco, Bonaventura Clotet, Nuria Prat, Marta Massanella, on behalf of the CoronAVI@S and the KING cohort extension studies

## Abstract

**Background:** SARS-CoV-2 vaccination is the most effective strategy to protect elders living in long-term care facilities (LTCF) against severe COVID-19, but primary vaccine responses are less effective in older adults. Here, we characterized the humoral responses following 3 months after mRNA/BNT162b2 vaccine in institutionalized elders.

**Methods:** Plasma levels of specific SARS-CoV-2 total IgG, IgM and IgA antibodies were measured before and 3 months after vaccination in elders living in LTCF. Neutralization capacity was assessed in a pseudovirus neutralization assay against WH1 (original) and B.1.617.2/Delta variants. A group of younger adults was used as reference group.

**Results:** Three months after vaccination, uninfected-elders presented reduced specific SARS-CoV-2 IgG levels and significantly lower neutralization capacity against the WH1 and Delta virus compared to vaccinated uninfected younger individuals. In contrast, COVID-19 recovered elders showed significantly higher specific SARS-CoV-2 IgG levels after vaccination than younger counterparts, while showing similar neutralization activity against WH1 virus and increased neutralization capacity against Delta variant. Despite previously infected elders elicit potent cross-reactive immune responses similarly to younger individuals, higher quantities of specific SARS-CoV-2 IgG antibodies are required to reach the same neutralization levels.

**Conclusions:** While hybrid immunity seems to be active in previously infected elders after three months from mRNA/BNT162b2 vaccination, humoral immune responses are diminished in COVID-19 uninfected vaccinated residents living in LTCF. These results suggests that a vaccine booster dose should be prioritized for this particularly vulnerable population.

**Word summary:** While previously infected and vaccinated elders living in LTCF had comparable neutralizing antibody levels to younger individuals, vaccinated uninfected-residents showed limited neutralization capacity against both original and delta variants. Hybrid immunity seems to be active in elders and can be relevant to design vaccine boosting campaigns.

## Introduction

Older adults have been disproportionately affected by the Coronavirus Disease 2019 (COVID-19) pandemic. Elders are more prone to a severe infection and over 95% of COVID-19-related mortality occurred in the population over 60 years of age [1]. Among them, residents from long-term care facilities (LTCF), who live in a congregate setting (with increased risk of transmission and infection) showed higher mortality rates than the general population of the same age [2,3]. Therefore, vaccination of residents from LTCF against SARS-CoV-2 has been a priority in most countries. SARS-CoV-2 vaccines are currently widely available in Europe, but we are still facing several months of continuous new infections and new circulating variants of concern (VOC). Indeed, virus outbreaks have been described in nursing homes during summer 2021 with B.1.617.2/Delta variant, despite the high vaccination rates among residents and healthcare professionals [4].

Currently available COVID-19 vaccines have shown to be safe and are able to generate an effective humoral and cellular immunity [5–8], but these studies have been performed including mainly young or middle-age healthy adults. Limited studies have evaluated the immune responses generated in elders living in LTCF [9–13], and most of them studied short term immune responses (several weeks after vaccination). Ageing is associated with an immunosenescent phenotype characterized by a progressive increase of a proinflammatory state and, a diminished immune response to pathogens and vaccines [14]. Therefore, there is an urgent need to determine the quality and the duration of immune responses of the elderly population, which could be very useful for designing specific SARS-CoV-2 vaccination calendars adapted to their immune needs.

To better understand how anti-SARS-CoV-2 humoral responses is triggered in the elderly population, we performed a prospective study to evaluate the anti-SARS-CoV-2 humoral response elicited after COVID-19 vaccination in both residents living in LTCF that have recovered from a previous SARS-CoV-2 infection (N=82) and SARS-CoV-2-uninfected individuals (N=16). The humoral response against SARS-CoV-2 was evaluated before and after 3 months of administration of the BNT162b2 mRNA COVID-19 vaccine (Pfizer-BioNTech). Humoral responses of elders were compared to a younger population, and a functional neutralization assay was performed against the WH1 virus and the Delta variant.

## Material and Methods

### Cohorts’ description

A total of 98 residents from three LTCF from the Northern are of Barcelona (Spain) were included in the prospective observational CoronAVI@S study (Table 1). Plasma samples were collected after 6 months from LTCF outbreaks (September-November 2020, pre-vaccine samples, N=98) and 3 months after complete vaccine schedule (post-vaccine samples, April-May 2021, N=91). Participants were recruited irrespective of their SARS-CoV-2 infection status. SARS-CoV-2 serology was performed to all collected pre-vaccine samples to determine their infection history. Elders were sub-classified in the infected and uninfected groups, depending on the PCR and serology status prior to vaccination (Supplementary Figure 1). Both study groups were vaccinated with BNT16b2 mRNA vaccine (Pfizer-BioNTech) from January to February 2021 according to the National COVID-19 vaccination strategy in Spain. An additional sample was collected a median of 2.8 months post-vaccination. We performed an additional serology test against NP-protein in uninfected residents after vaccination to confirm their SARS-CoV-2-naïve status (data not shown).

**Table 1:** Descriptive table of CoronAVI@S participants.

|  | Previously SARS-CoV-2 Infected group | Uninfected group | *P-value* |
| --- | --- | --- | --- |
|  | N= 82 | N=16 |  |
| Age, years, median [IQR] | 87 [81-90] | 79 [74-90] | 0.26 |
| Female, n (%) | 66 (80) | 8 (50) | **0.021** |
| GMA<sup>2</sup>, n (%) |  |  | 0.11 |
| 2 | 4 (5) | 3 (20) |  |
| 3 | 38 (46) | 6 (40) |  |
| 4 | 41 (49) | 6 (40) |  |
| **Visit pre-vaccine** | **N=82** | **N=16** |  |
| PCR positive, n (%) | 61 (73) | 0 (0) | **<0.0001** |
| Time from PCR positive to sample extraction, months, median [IQR] | 6.4 [6-6.9] | NA | NA |
| Detection of SARS-CoV-2 specific antibodies, n (%) | 81 (98)<sup>1</sup> | 0 (0) | **<0.0001** |
| Medical assistance, n (%) | 22 (27) | NA |  |
| **Visit post-vaccine** | **N=76** | **N=15** |  |
| Vaccine received |  |  |  |
| BNT16b2 (2 doses), n (%) | 76 (100) | 15 (100) | 1.0 |
| Time between vaccine doses, days, median [IQR] | 21 [21-23] | 21 [21-26] | 0.14 |
| Time between PCR positive and 1st vaccine dose, months, median [IQR] | 9.3 [9.1-9.6] | NA |  |
| Time from complete vaccine schedule to sample extraction, months, median [IQR] | 2.4 [2.36-2.95] | 2.83 [2.6-3.06] | 0.05 |
| PCR positive after vaccination, n (%) | 0 (0) | 1 (7) |  |

<sup>1</sup>One individual did not seroconvert despite Positive PCR
<sup>2</sup>GMA: Stratum of Adjusted Morbidity ranging from 1 to 4 according to the number of comorbidities and their need of health care

For comparing our data with a younger population with less comorbidities, we selected a total of 110 infected participants (Table 2) and 51 individuals (32 infected and 16 uninfected individuals, Table 3), for pre-vaccine and post vaccine comparisons, respectively from the *KING cohort extension*. The age range of selected participants were between 22 and 64 years. Blood samples of pre-vaccine were matched with CoronAVI@S study by time post-SARS-CoV-2 infection (6 months). Post-vaccine samples were selected among individuals vaccinated also with BNT16b2 (Pfizer-BioNTech) and blood samples were obtained a median of 3 months after complete vaccination schedule.

**Table 2:** Descriptive table of younger and older participants pre-vaccine.

|  | CoronAVI@S |  | KING Cohort extension | *P-value* |
| --- | --- | --- | --- | --- |
|  | Infected<br>N= 82 | Uninfected<br>N=16 | Infected<br>n=110 | Infected<br>*Younger vs. Older* |
| Age, years, median [IQR] | 87 [81-90] | 79 [74-90] | 48[40-55] | **<0.001** |
| Age, years, n (%) |  |  |  |  |
| <45 | 0 (0) | 0 (0) | 50 (45) |  |
| 45-65 | 0 (0) | 0 (0) | 60 (55) |  |
| 65-85 | 37 (45) | 10 (63) | 0 (0) |  |
| >85 | 45 (55) | 6 (37) | 0 (0) |  |
| Female, n (%) | 66 (80) | 8 (50) | 64 (58) | **0.0011** |
| History of SARS-CoV-2 infection, n (%) | 82 (100) | 0 (100) | 110 (100) | **<0.001** |
| ***Visit pre-vaccine*** |  |  |  |  |
| Time from PCR positive/symptoms to sample extraction, months, median [IQR] | 6.5 [6.0-6.9] | NA | 6.6 [7.2-6.1] | **0.04** |
| Detection of SARS-CoV-2 specific antibodies, n (%) | 82 (99) | 0(0) | 106 (96) | 0.39 |
| Medical assistance, n (%) | 22 (27) |  | 45 (41) | **0.0001** |

**Table 3:** Descriptive table of younger and older participants pre-vaccine.

|  | CoronAVI@S |  | KING Cohort extension |  | *P-values between younger and older* |  |
| --- | --- | --- | --- | --- | --- | --- |
|  | Infected<br>n= 76 | Uninfected<br>n= 15 | Infected<br>n=32 | Uninfected<br>n=19 | Infected | Uninfected |
| Age, years, median [IQR] | 87 [81-90] | 79 [74-90] | 43 [38-50] | 39 [28-48] | **<0.001** | **<0.001** |
| Age, years, n (%) |  |  |  |  |  |  |
| ≤45 | 0 (0) | 0 (0) | 19 (59) | 12 (63) |  |  |
| 46-65 | 0 (0) | 0 (0) | 13 (41) | 7 (37) |  |  |
| 65-85 | 33 (43) | 10 (67) | 0 (0) | 0 (0) |  |  |
| >85 | 43 (57) | 5 (33) | 0 (0) | 0 (0) |  |  |
| Female, n (%) | 62 (82) | 8 (53) | 22 (69) | 14 (74) | 0.20 | 0.28 |
| History of SARS-CoV-2 infection, n (%) | 76 (100) | (1)<sup>1</sup> | 32 (100) | NA | 1.0 | NA |
| Medical assistance, n (%) | 21 (27) | NA | 2 (6) | NA | **0.02** | NA |
| Vaccine received, n (%) |  |  |  |  |  |  |
| BNT16b2, 1 doses | 76 (100) | 15 (100) | 32 (100) | 19 (100) | 1.0 | 1.0 |
| BNT16b2, 2 doses | 76 (100) | 15 (100) | 30 (94) | 19 (100) | 0.08 | 1.0 |
| Time between vaccine doses, days, median [IQR] | 21 [21-23] | 21 [23-26] | 22 [21-22] | 22 [21-22] | 0.34 | 0.23 |
| Time between PCR positive/symptoms and complete vaccine schedule, months, median [IQR] | 12.9 [12.7-13.3] | NA | 10.6 [10.4-11.6] | NA | **<0.001** | NA |
| Time from complete vaccine schedule to sample extraction, months, median [IQR] | 2.4 [2.36-2.95] | 2.83 [2.6-3.06] | 2.04 [2.87-3.07] | 3.1 [3.01-3.37] | 0.53 | **<0.001** |

<sup>1</sup>One infection post-vaccine

The CoronAVI@S study and the KING cohort extension were approved by the Ethics Committee Boards from the Institut Universitari d’Investigació en Atenció Primària Jordi Gol (IDIAP-JGol) and the Hospital Universitari Germans Trias i Pujol (HUGTIP), respectively (IDIAP-JGol /20-116P and HUGTiP/PI-20-217). All participants provided written informed consent before inclusion.

### Determination of anti-SARS-CoV-2 antibodies

The presence of anti-SARS-CoV-2 antibodies against S2+RBD or NP in plasma samples was evaluated using an in-house developed sandwich-ELISA, as previously described [15]. The specific signal for each antigen was calculated after subtracting the background signal obtained for each sample in antigen-free wells. Values are plotted into the standard curve. Standard curve was calculated by plotting and fitting the log of standard dilution (in arbitrary units) vs. response to a 4-parameter equation in Prism 8.4.3 (GraphPad Software). See supplemental material.

#### Pseudovirus neutralization assay

Neutralization assay was performed using SARS-CoV-2.SctΔ19 WH1 and B.1.617.2/Delta pseudovirus as previously described [16,17]. The lower limit of detection was 60 and the upper limit was 14,580 (reciprocal dilution). The ID50 (the reciprocal dilution inhibiting 50% of the infection) was calculated by plotting and fitting the log of plasma dilution vs. response to a 4-parameter equation. See supplemental material.

### Statistics

Continuous variables were described using medians and the interquartile range (IQR, defined by the 25th and 75th percentiles), whereas categorical factors were reported as percentages over available data. Quantitative variables were compared using the Mann–Whitney test and percentages using the chi-squared test for comparison between groups, Wilcolxon for paired tests and Kruskal–Wallis test for comparison between age ranges. Analyses were performed with Prism 9.1.2 (GraphPad Software).

## Results

### Characterization of older adults living in LTCF

A total of 98 residents from three LTCF in the Northern area of Barcelona (Spain) were included in the prospective observational CoronAVI@S study. All participants were >65 years, with 90% of the individuals belonging to the moderate to high risk groups using GMA (Adjusted Morbidity Group) [18]. Most common chronic pathologies included hypertension (62%), arthritis (40%), dementia (38%) or diabetes (33%). Individuals were tested by RT-PCR during LTCF outbreak from March to June 2020.

Serologic and PCR tests before vaccination revealed that 84% resident included in the study suffered from previous SARS-CoV-2 infection. Previously infected residents (n=82) had a median age of 87 years and were 80% female, whereas uninfected residents before vaccination (n=16) had a median age of 79 years and were 50% female. Additional characteristics of the participants at enrolment are shown in Table 1.

We took advantage of the national vaccination plan in LTCF to evaluate the immune responses generated by the BNT16b2 (Pfizer-BioNTech). Only one uninfected resident was infected with SARS-CoV-2 after vaccination (confirmed by NP-serology and RT-PCR), developing a mild COVID-19. This individual is shown in red in all graphs, and excluded from statistical analysis.

### Humoral responses after 3 months from vaccination

We first evaluated the impact of vaccination in the levels of circulating specific SARS-CoV-2 antibodies in elders living in LTCF. The levels of SARS-CoV-2 specific IgG, IgA and IgM antibodies were significantly higher in individuals who were infected prior vaccination compared to their uninfected counterparts (p<0.0001 in all cases, Figure 1A). There was a significant increase of all isotypes of immunoglobulins between pre- and post-vaccine samples in infected elders (Supplementary 2A), while uninfected elders maintained detectable levels of specific SARS-CoV-2 IgG and IgA antibodies, but no IgM, three months after vaccination (Supplementary 2B). Of note, the individual who got infected after vaccination showed similar levels of specific SARS-CoV-2 antibodies than the infected and vaccinated group. We then evaluated the levels of SARS-CoV-2 specific IgG antibodies induced after vaccination across age groups. We observed higher levels of IgG antibodies in older groups (≥65 years) previously infected compared to younger individuals (Figure 1B, Anova p<0.001, and r= 0.41, p<0.001, Supplementary Figure 2C). To determine if the higher levels of virus-specific IgG antibodies in infected elders after vaccination was related to the vaccine or to the natural infection, we evaluated specific IgG antibodies before vaccination across ages (Supplementary Figure 2D). We determined that infected residents after six months from infection (and prior vaccination) showed already higher levels of SARS-CoV-2 IgG antibodies compared to the younger population (Anova p<0.001). In contrast, the levels of circulating specific SARS-CoV-2 IgG antibodies in uninfected vaccinated subjects tended to decrease across age groups (Anova p =0.06, Figure 1B), and negatively correlated with age (r=-0.72, p<0.001, Supplementary Figure 2C). The levels of SARS-CoV-2 specific IgG antibodies were significantly decreased already in individuals between 45 and 64 years old when comparing infected and uninfected group prior vaccination (p=0.007), and this difference was maintained in the older age groups (p<0.001 in all cases, Figure 1B). These results highlighted some differences in antibody production between infected and uninfected individuals, being only the uninfected younger population able to produce similar levels of SARS-CoV-2 antibodies to their infected counterparts.

**Figure 1:**
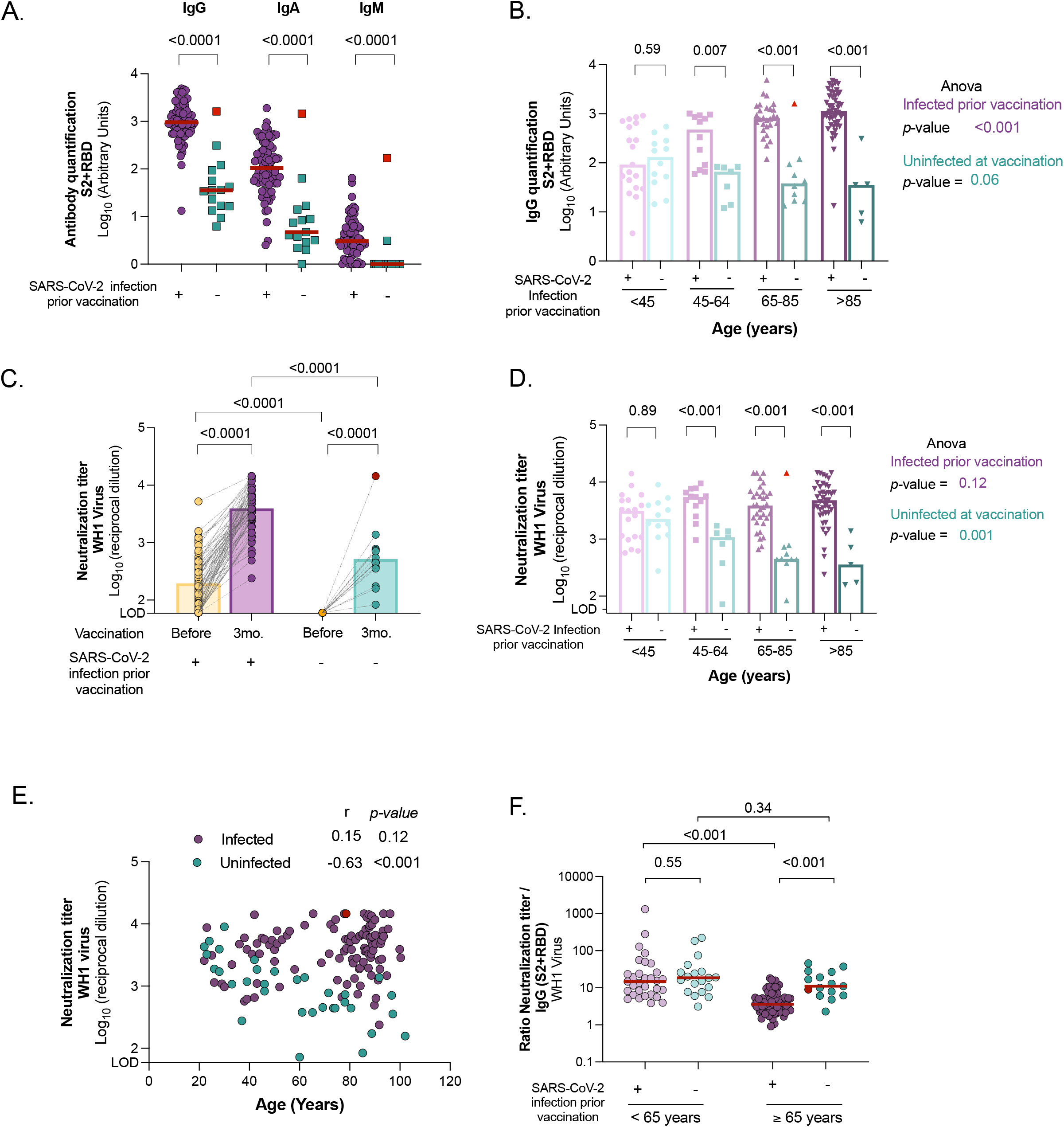
Comparison of humoral response and neutralizing activity between uninfected and infected individuals at different ages after three months from BNT162b2 mRNA COVID-19 vaccine. <u>Panel A</u>: Levels of specific SARS-CoV-2 immunoglobulins (IgG, IgA and IgM) against S2+RBD proteins quantified in plasma from uninfected and infected elders by ELISA. <u>Panel B</u>: SARS-CoV-2 specific IgG antibody levels (against S2+RBD proteins) after vaccination across ages in infected and uninfected participants. <u>Panel C</u>: Neutralizing activity against WH1 virus before and after three months of vaccine administration in infected and uninfected elders living in LTCF. <u>Panel D:</u> Neutralizing activity against WH1 after vaccination across ages in infected and uninfected participants. <u>Panel E:</u> Correlation of neutralizing activity after vaccination with age in participants infected and uninfected. Correlation coefficient and p-values were obtained from Spearman correlation. <u>Panel F:</u> Ratio of plasma neutralization titer per total SARS-CoV-2 IgG antibodies in younger and older individuals, sub-grouped by previous SARS-CoV-2 infection history. Median values are indicated; P-values were obtained from Mann–Whitney test for comparison between groups (Panels A, B, C, D and F), Wilcolxon for paired tests (panel C) and Kruskal–Wallis test for comparison between ranges of age for infected and uninfected groups (Panel B and D). In all panels, uninfected and infected individuals at vaccination are indicated in turquoise and purple, respectively. Uninfected resident who got infected after vaccinations is indicated in red and was excluded from the statistical analysis.

### Neutralization capacity against original WH1 virus

All residents were assayed for their plasma neutralization capacity against the Wuhan-Hu-1 (WH1) sequence in a validated pseudovirus assay [16,17]. SARS-CoV-2 vaccination increased the titers of antibodies with neutralizing capacity in infected and uninfected elders (p<0.0001 in both cases, Figure 1C), being the neutralization titers significantly higher in individuals with a previous history of SARS-CoV-2 infection compared to the uninfected counterparts (p<0.0001). For the infected population, there was a median fold increase of 18.7 [9.3-44.8] between pre-vaccine sample compared to post-vaccination sample. Of note, the resident who was infected after vaccination showed similar titers of neutralizing antibodies as the infected group after vaccination. We then compared the plasma neutralization capacity between residents and younger individuals, observing that previously infected subjects showed similar levels of neutralization independently of age (Anova p=0.12, Figure 1D and Supplementary Figure 2E). We hypothesized that the neutralization response after natural infection reached a similar plateau in recovered elders than younger subjects, therefore similar levels of neutralization were reached in both groups after vaccination. For that, we compared the plasma neutralization capacity before vaccination between older and younger individuals after six months from SARS-CoV-2 infection. Surprisingly, the neutralization titers were significantly lower in elders compared to younger after six months from infection (Anova p<0.001, Supplementary Figure 2F). These differences were not related to the severity of COVID-19, since no differences were observed in elders with mild or severe disease (Supplementary Figure 2G). These results suggest a greater boost of humoral responses in infected elders after vaccination compared to the younger group. In contrast, uninfected vaccinated subgroups showed sequential decrease of neutralization titers across ages (Anova p=0.001 Figure 1D and Supplementary Figure 2H). Indeed, age was only negatively correlated with plasma neutralization capacity in the uninfected group (r=-0.63, p<0.001; Figure 1E). Overall, these results suggest that uninfected older adults show lower titers of neutralizing antibodies and might be at higher risk of infection than previously infected individuals of the same age.

We then evaluated the proportion of effective neutralizing antibodies among the total SARS-CoV-2 specific IgG, calculated as the ratio of plasma neutralization titer to total SARS-CoV-2 IgG antibodies. We observed a reduction of this ratio in infected elders compared to their younger counterparts (Figure 1F, p<0.001), suggesting low functionality of antibodies in elders, where high levels of circulating SARS-CoV-2 IgG are required to reach protection. In contrast, no differences were found between uninfected and vaccinated younger and older adults (Figure 1C). These results suggest that mRNA vaccine induce effective neutralizing antibodies in elders, but at lower levels.

### Neutralization capacity against Delta variant

Given our observation that uninfected LTCF residents aged ≥65 years had lower neutralization responses, following complete vaccination schedule than younger individuals, we hypothesized that this particular group could experience lower neutralizing responses against the VOC B.1.617.2/Delta, which has been the main variant circulating in Spain since July 2021. Indeed, we found a significant decrease in the plasma neutralization capacity of Delta VOC compared to WH1 in all elders and younger adults, independently of their previous infection status (p<0.001 in both cases, Figure 2A and Supplementary Figure 3 A-C). We then examined plasma neutralization capacity by age group against Delta variant. Similarly, to the original virus (WH1), we found a progressive decrease of the levels of neutralization across ages in vaccinated uninfected individuals (Figure 2B and Supplementary Figure 3D, Anova P=0.005). Remarkably, neutralization capacity against Delta variant was barely detected in uninfected vaccinated individuals older than 65 years (except for the resident who got infected after vaccination). Surprisingly, neutralization capacity against Delta variant increased significantly across ages in individuals with prior SARS-CoV-2 infection (Figure 2B and Supplementary Figure 3E, Anova P<0.001). Next, we compared cross-neutralization capacities by determining fold change ratios between Delta and WH1 (reference spike) virus, as a measure of the loss of neutralizing capacity (a ratio <1 indicates a better neutralization of the reference spike of WH1 in comparison to the delta, and vice versa). Median fold-change of all groups were <1, highlighting the higher difficulty to neutralize Delta variant compared to the original one (Figure 2C). Among all groups, previously infected elders after vaccination showed significantly higher ratios compared to all other groups, demonstrating the better cross-neutralization of the Delta variant.

**Figure 2:**
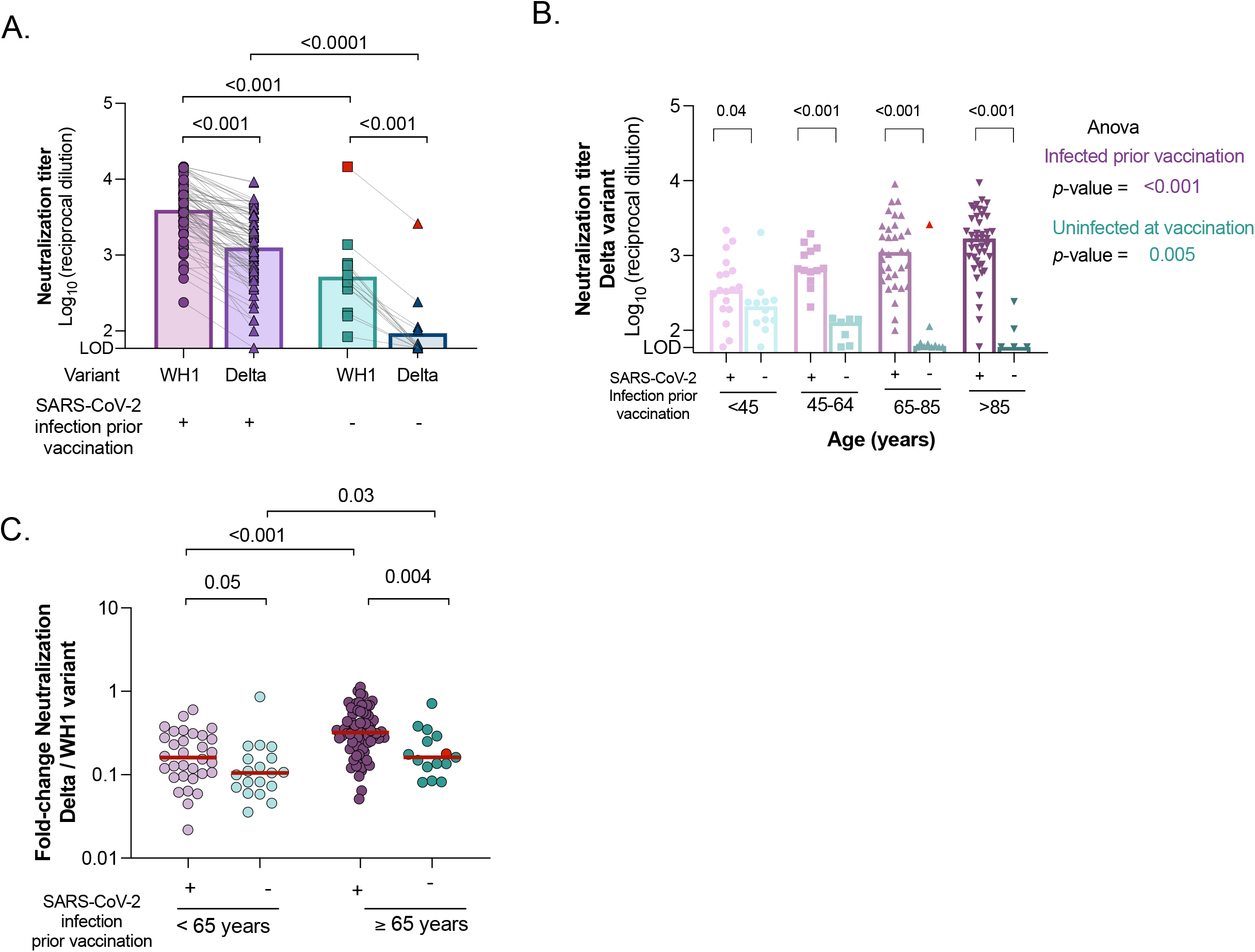
Neutralizing activity against B.1.617.2/Delta variant between uninfected and infected individuals at different ages after three months from BNT162b2 mRNA Covid-19 vaccine. <u>Panel</u> <u>A</u>: Comparison of plasma neutralizing activity against WH1 and B.1.617.2/Delta variant in uninfected and infected resident elders after vaccination. <u>Panel B:</u> Neutralizing activity against Delta variant after vaccination across ages in infected and uninfected, younger and older individuals, sub-grouped by previous SARS-CoV-2 infection history. <u>Panel C</u>: Fold change ratios between neutralization titers against Delta variant and WH1 (reference spike) virus in infected and uninfected individuals, subgroup by age (<65 years and ≥65 years). Median values are indicated. P-values were obtained from Mann–Whitney test for comparison between groups (all panels), Wilcolxon for paired tests (Panel A and C) and Kruskal–Wallis test for each group (Panel B). In all panels, uninfected and infected individuals at vaccination are indicated in turquoise and purple, respectively. Uninfected resident who got infected after vaccinations is indicated in red was excluded from the statistical analysis.

## Discussion

LTCF residents have been prioritised for COVID-19 vaccination in the majority of countries due to their increased risk of morbidity and mortality. Although the effectiveness of COVID-19 vaccines is very high, SARS-CoV-2 infections among fully vaccinated individuals (i.e., breakthrough infections) are expected, especially with the appearance of VOC with higher transmissibility and immune resistance, compared to the original WH1 virus. The reduced ability of elders to elicit effective immune responses to vaccine and pathogens, place this population once again as one of the most vulnerable populations that need to be carefully analysed.

In this study, we have demonstrated that uninfected residents after three months from COVID-19 vaccination showed limited neutralization responses compared to the younger uninfected individuals, putting the uninfected elder population at higher risk. Vaccination with two doses of BNT162b2 of uninfected LTCF residents has been shown to reduce the risk of infection by 90%, and more than 95% in COVID-19 related hospital admission and mortality for up to 5 months [19]. These results combined with ours suggest that uninfected elders may develop a memory response, which protect them in case of a new infection. However, the low levels of circulating neutralizing antibodies after three months highlight the need of further studies to assess the long-term effectiveness of SARS-CoV-2 vaccines, and may suggest that this specific subgroup may benefit from a boost vaccine [20]. Of note, the resident who got infected after vaccination in our study, clearly show a boost of neutralization responses to the levels of the infected groups after vaccination and developed a mild COVID-19.

Importantly, previously infected elders have developed a particularly potent immune responses to COVID-19 vaccines, similar to younger individuals [10,12,13,21]. Our results suggest that the combination of natural and vaccine-generated immunity, known as hybrid immunity [22], elicit a larger than expected immune responses even in elders, despite a high degree of immunosenescence. It has been recently shown that hybrid immunity improves B cells and antibodies against SARS-CoV-2 variants [23]. Elders show higher capacity to neutralize more resistant variants (i.e., B.1.617.2/Delta) than younger individuals, being the hybrid immunity a benefit for this recovered population.

Our results demonstrate that natural infection (pre-vaccine) generates higher virus-specific IgG antibodies in elders compared to the younger population. In contrast, the vaccination of uninfected elders generates lower SARS-CoV-2-specific IgG antibodies, as other studies have already suggested [9,10,12,13], and these antibodies show lower neutralization capacity compared to the younger population. On the other hand, previously infected elders after vaccine show high levels of total SARS-CoV-2 IgG compared to the younger counterparts, with similar levels of neutralizing antibodies. Therefore, the measurement of SARS-CoV-2-specific IgG antibodies might not be a good surrogate marker of protection in elders and functional assays, such as virus/pseudovirus based neutralization tests, are required.

Some studies have shown reduced levels of neutralizing titters in uninfected elders after 2 to 3 weeks from BNT162b2 vaccination compared to the younger population [11,12], suggesting already a limited initial immune response of older individuals after vaccine administration. In addition, Collier *et al*. observed that serum from elders after three weeks from vaccination showed lower, but detectable neutralization capacity to different VOC (alpha, beta or gamma) than younger population [11]. Our results showed that plasma from uninfected LTCF residents were unable to neutralize Delta variant after three months from vaccination. This could be due to the more resistant nature of the SARS-CoV-2 Delta variant to be inhibited by specific antibodies [24] and/or the lower durability of neutralizing antibodies in elders compared to younger individuals (3 weeks *vs*. 3 months) [9]. Nonetheless, the low levels of neutralizing antibodies of uninfected elders against the original virus (WH1) after 3 months from vaccination, and even lower against Delta variant, may suggest that this specific population may require a third dose of the SARS-CoV-2 vaccine to boost their humoral response. Indeed, a booster vaccine (3^rd^ dose) in >60 years population from Israel have demonstrated a diminution of confirmed COVID-19 and severe illness [20,25], supporting our recommendation. On the other hand, it has been demonstrated that mRNA vaccination boosts cross-variant antibodies elicited by natural infection in the general population [26,27]. Our results are in line with these findings and clearly reveal that elders previously infected developed the same cross-neutralization capacity against Delta variant than the younger population. Therefore, previously infected elders would not benefit from an additional vaccine dose at this time. Additional studies might be required to assess the durability of plasma neutralization capacity in infected elders compared to the younger population.

Even though an assessment of the immune response several months after vaccination has been performed, our study has some limitations. First, the relatively small number of participants, especially for the uninfected elder group, which was unexpectedly low. While 64% of the cohort tested positive by PCR in massive tests screenings during outbreaks, SARS-CoV-2 serologic test revealed that 84% of residents suffered from COVID-19. In addition, we did not assess specific SARS-CoV-2 cellular responses before and after vaccination, which could also be used as a correlate of protection [28].

Further follow-up of the immune responses, as well as registration of breakthrough infections in COVID-19-uninfected residents from LTCF will allow to define the immune correlate of protection in this frail and vulnerable population. To this purpose, determination of the quality of humoral immune response, using functional neutralization assays, is necessary in the population older than 65 years. Taken together, our results suggest that only the uninfected-residents, who do not develop adequate immune responses, will clearly benefit from a booster vaccine dose; an adapted vaccination calendar would be necessary to respond to the immune needs of this vulnerable population. Importantly, hybrid immunity seems to be active in elders and can be relevant to design vaccine boosting campaigns.

## Supporting information

All Supplemental Figures

## Data Availability

All data produced in the present study are available upon reasonable request to the authors

## Declarations

### Competing interests

JB and JC reports personal fees from Albajuna Therapeutics, outside the submitted work.

### Funding

This work was partially funded by Gloria Soler Foundation, Grifols, the Departament de Salut of the Generalitat de Catalunya (grant DSL0016 to JB and Grant DSL015 to JC), the Spanish Health Institute Carlos III (Grant PI17/01518 and PI18/01332 to JC) and the crowdfunding initiatives “https://www.yomecorono.com“, BonPreu/Esclat and Correos. MT and CAN were supported by the early-stage research staff Fellowship (FI21-B00327 and FI20-B00742) from the Catalan Agency for Management of University and Research Grants (AGAUR) and the European Social Fund. MT was then supported by a doctoral fellowship from the Departament de Salut from Generalitat de Catalunya (SLT017/20/000095). EP was supported by a doctoral grant from ANID, Chile: Grant 72180406.

### Author’s Contributors

MM, NP and BC conceived the study. MT, TP, CA-N and JC determined and analysed anti-SARS-CoV-2 specific antibodies. MT, EP, BT, SM, CR and JB determined and analysed neutralization activity. DP, MI, NM, JMB, NP, EG, AC, AM, RT, MF and LM contribute to clinical management and sample collection. MT and MM analysed the data and prepared the figures. MM and NP coordinated the study, collected all clinical and laboratory data and wrote the manuscript. All authors discussed the results and approved the manuscript.

## Acknowledgements

We are deeply grateful to all participants and their families, to the LTCF who participated in this study. We also thank the technical staff of Direcció d’Atenció Primària de la Metropolitana Nord for sample collection in LTCF (S. Reyes Carrión, N. Salarich Solà, A. Vidal, R. Alvarez Viñallonga, J. Tornero, E. Vilamala, C. Suarez, T. Gonzalo, L. Perez, D. Sans, A. Blancas Loras, A. Garcia Archer, J. Borràs, S. Cervelló) and staff of IrsiCaixa for sample processing (L. Ruiz, R. Ayen, L. Gomez, C. Ramirez, M. Martinez, T Puig). We thank “CERCA Programme/Generalitat de Catalunya for institutional support and the Foundation Dormeur.

## Figure legends

<u>**Supplementary Figure 1**</u>: **Flowchart Coronavi@s study**, including 98 elders living in LTCF. Number of samples analysed at each time point are indicated.

<u>**Supplementary Figure 2**</u>: **Comparison of humoral response and neutralizing activity between uninfected and infected individuals at different ages before and after three months from BNT162b2 mRNA Covid-19 vaccine.** Levels of specific SARS-CoV-2 immunoglobulins (IgG, IgA and IgM) against S2+RBD proteins quantified in plasma from infected (Panel A) and uninfected (Panel B) elders by ELISA, before and after vaccination. <u>Panel C</u>: Correlation of the levels of specific SARS-CoV2 IgG antibodies (S2+RBD) after vaccination with age in participants infected and uninfected. Correlation coefficient and p-values were obtained from Spearman correlation. <u>Panel D:</u> SARS-CoV-2 specific IgG antibody levels (against S2+RBD proteins) after 6 months from infection (and before vaccination) across ages in infected participants. Plasma neutralizing activity against WH1 variant across ages in infected (Panel E) or uninfected (Panel H). <u>Panel F</u>: Plasma neutralizing activity against WH1 variant across ages in participants after six months from symptoms onset (and before vaccination); individuals were sub-grouped according to their medical assistance requirements (Panel F). Median values are indicated. P-values were obtained from Mann–Whitney test for comparison between groups (Panel F), Wilcolxon for paired tests (panel A and B), Kruskal–Wallis test for each group (Panel D, E, F, G H). In all panels, uninfected and infected individuals at vaccination are indicated in turquoise and purple, respectively. Uninfected resident who got infected after vaccinations is indicated in red was excluded from the statistical analysis.

<u>**Supplementary Figure 3**</u>: **Neutralizing activity against B.1.617.2/Delta variant between uninfected and infected individuals at different ages after three months from BNT162b2 mRNA Covid-19 vaccine.** <u>Panel A:</u> Comparison of plasma neutralization capacity after vaccination against WH1 virus (original) and Delta variant in infected and uninfected individuals. Comparison of plasma neutralization capacity after vaccination against WH1 virus (original) and Delta variant across ages in Uninfected (Panel B) and infected individuals (Panel C). Neutralizing activity against Delta variant after vaccination across ages in uninfected (Panel D) and infected (Panel E). In all panels, median values are indicated and P-values were obtained from Mann-Whitney test (Panel A) and Wilcolxon for paired tests (Panel B and C) and Kruskal– Wallis test for each group (Panel D and E).

## Notes

### Funding Statement

This study was funded by Gloria Soler Foundation, Grifols, the Departament de Salut of the Generalitat de Catalunya (grant DSL0016 to JB and Grant DSL015 to JC), the Spanish Health Institute Carlos III (Grant PI17/01518 and PI18/01332 to JC) and the crowdfunding initiatives https://www.yomecorono.com, BonPreu/Esclat and Correos. MT and CAN were supported by the early-stage research staff Fellowship (FI21-B00327 and FI20-B00742) from the Catalan Agency for Management of University and Research Grants (AGAUR) and the European Social Fund. MT was then supported by a doctoral fellowship from the Departament de Salut from Generalitat de Catalunya (SLT017/20/000095). EP was supported by a doctoral grant from ANID, Chile: Grant 72180406.

### Author Declarations

Ethics committe of IDIAP Jordi Gol (granted by The Office for Human Research Protections (OHRP) - US Department of Health & Human Services Office of the Secretary office of Public Health & Science office for Human Research Protection: IRB00005101 / IORG0004303 / FWA Number 00009235) gave ethical approval for this work. This study only uses Pseudo-anonymized data.

