## Supplementary material for "Limited immune responses after three months of BNT162b2 vaccine in SARS-CoV-2 uninfected elders living in long-term care facilities": All Supplemental Figures

Supplementary material and methods

#### Determination of anti-SARS-CoV-2 antibodies

Nunc MaxiSorp ELISA plates were coated overnight at 4ºC with 50ng/mL of capture antibody (anti-6xHis antibody, clone HIS.H8; ThermoFisher Scientific) at 2 μg/mL in PBS. After washing, plates were blocked for two hours at room temperature using PBS/1% of bovine serum albumin (BSA, Miltenyi biotech). Then, 50 µL of the following SARS-CoV-2 derived antigens diluted in in blocking buffer were added: Spike (S2) (0.9 µg/mL), receptor binding domain (RBD) (0.3 µg/mL) or nucleocapsid protein (NP) (1 µg/mL) (Sino Biologicals) and incubated overnight at 4ºC. Each plasma sample was evaluated in duplicated at dilution ranging from 1/100- 1/50000 in blocking buffer for each antigen. Diluted samples were incubated at room temperature for one hour. Antigen free wells were also assayed in parallel for each sample in the same plate to evaluate sample background. Serial dilutions of a positive plasma sample were used as standard. A pool of 10 SARS-CoV-2 negative plasma samples, collected before June 2019, were included as negative control. The following reagents were used as secondary antibodies: HRP conjugated (Fab)2 Goat anti-human IgG (Fc specific) (1/20,000), Goat anti-human IgM (1/10,000), and Goat anti-human IgA (alpha chain specific) (1/10,000) (all from Jackson Immunoresearch). Secondary antibodies were incubated for 30 minutes at room temperature. After washing, plates were revealed using o-Phenylenediamine dihydrochloride (OPD, Sigma Aldrich) and the enzymatic reaction was stopped with 4N of $H_2SO_4$ (Sigma Aldrich). The signal was analysed as the optical density (OD) at 492 nm with noise correction at 620 nm. The specific signal for each antigen was calculated after subtracting the background signal obtained for each sample in antigen-free wells. Values are plotted into the standard curve. Standard curve was calculated by plotting and fitting the log of standard dilution (in arbitrary units) vs. response to a 4-parameter equation in Prism 8.4.3 (GraphPad Software).

#### Pseudovirus neutralization assay.

SARS-CoV-2.SctΔ19 WH1 and B.1.617.2/Delta were generated (Geneart) from the full protein sequence of the original SARS-Cov-2 isolate Wuhan-Hu-1 (WH1) and the Delta variant (B.1.617.2) spike sequences respectively, with the deletion of the last 19 amino acids in Cterminal [15], human-codon optimized and inserted into pcDNA3.1(+). HIV reporter pseudoviruses expressing SARS-CoV-2 S protein and Luciferase were generated using the defective HIV plasmid pNL4-3.Luc.R-.E- obtained from the NIH AIDS Reagent Program [16]. Expi293F cells were transfected using ExpiFectamine293 Reagent (Thermo Fisher Scientific) with pNL4-3.Luc.R-.E- and SARS-CoV-2.SctΔ19 (WH1, B.1.617.2/Delta), at an 8:1 ratio, respectively. Control pseudoviruses were obtained by replacing the S protein expression plasmid with a VSV-G protein expression plasmid as reported [17]. Supernatants were harvested 48 h after transfection, filtered at 0.45 µm, frozen and titrated on HEK293T cells overexpressing WT human ACE-2 (Integral Molecular). Neutralization assays were performed in duplicate. Briefly, in Nunc 96-well cell culture plates (Thermo Fisher Scientific), 200 TCID50 of pseudovirus were preincubated with three-fold serial dilutions (1/60–1/14,580) of heat-inactivated plasma samples for 1 h at 37 ∘C. Then, $2 \times 10^4$ HEK293T/hACE2 cells treated with DEAE-Dextran (Sigma-Aldrich) were added. Results were read after 48 h using the EnSight Multimode Plate Reader and BriteLite Plus Luciferase reagent (Perkin Elmer, Waltham, MA, USA). The values were normalized, and the ID50 (the reciprocal dilution inhibiting 50% of the infection) was calculated by plotting and fitting the log of plasma dilution vs. response to a 4-parameter equation in Prism 8.4.3 (GraphPad Software). This neutralization assay had been previously validated in a large subset of samples [18,19]. The lower limit of detection was 60 and the upper limit was 14,580 (reciprocal dilution).

### Supplementary Figure 1

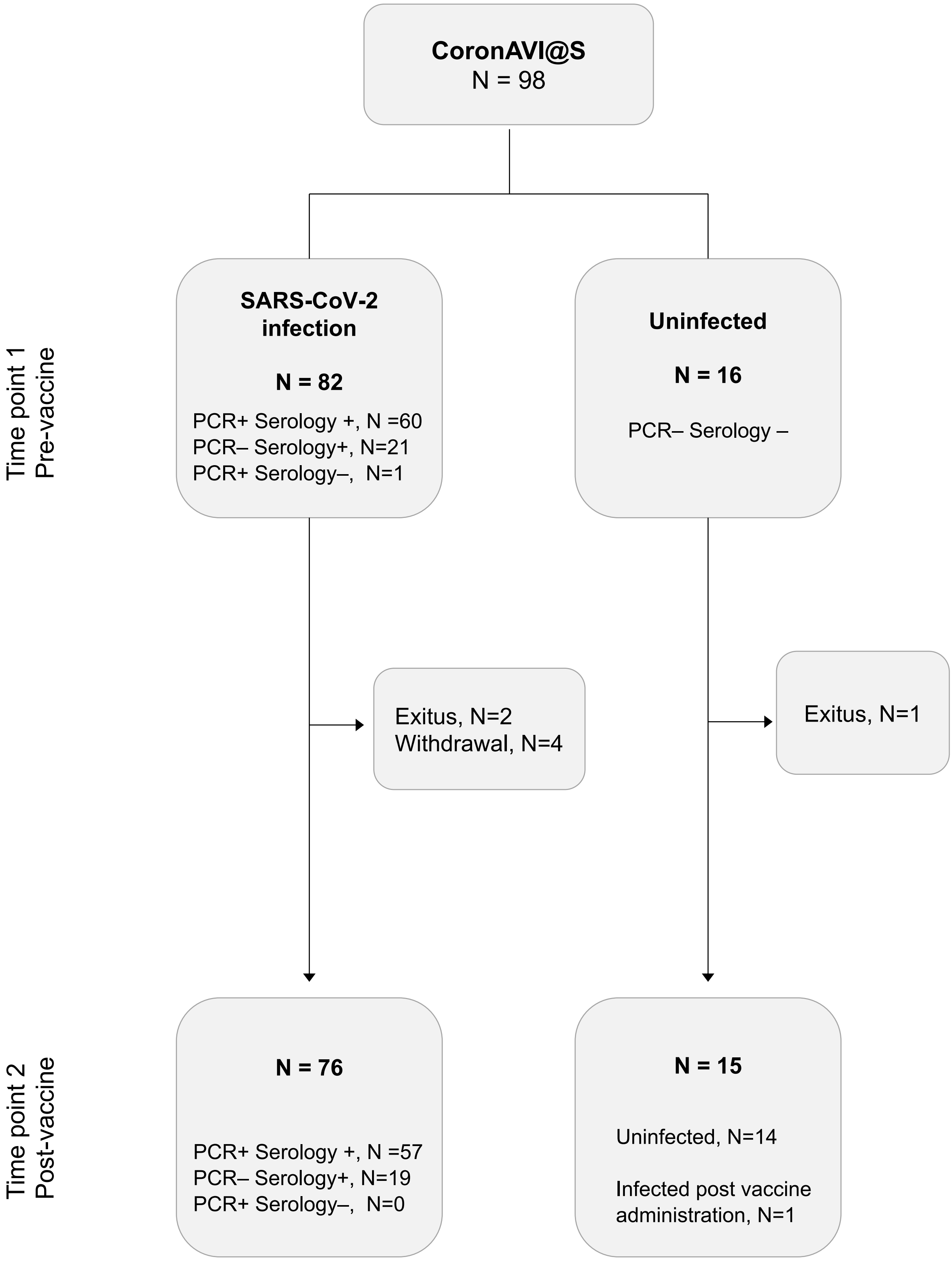

### Supplementary Figure 2

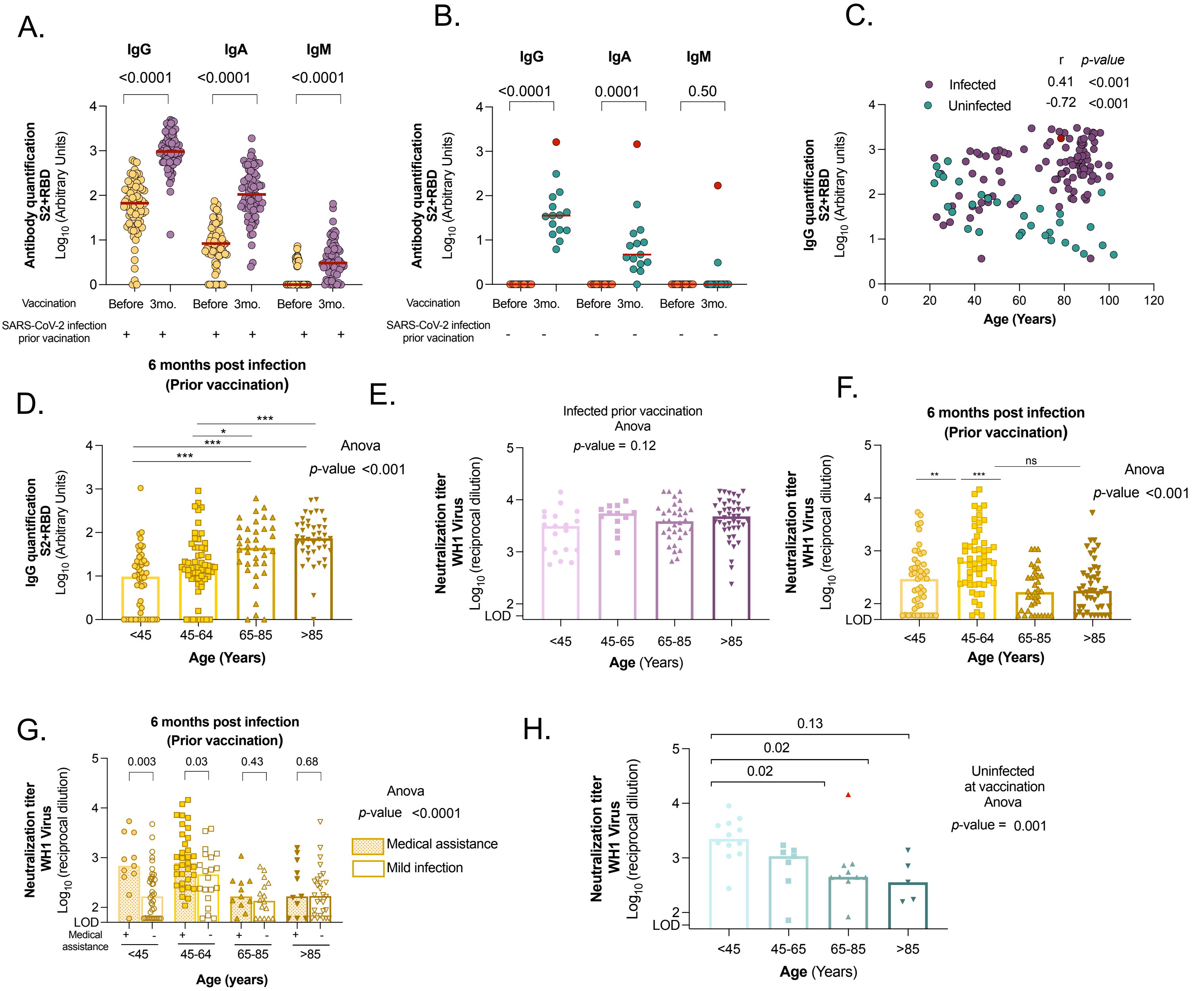

### Supplementary Figure 3

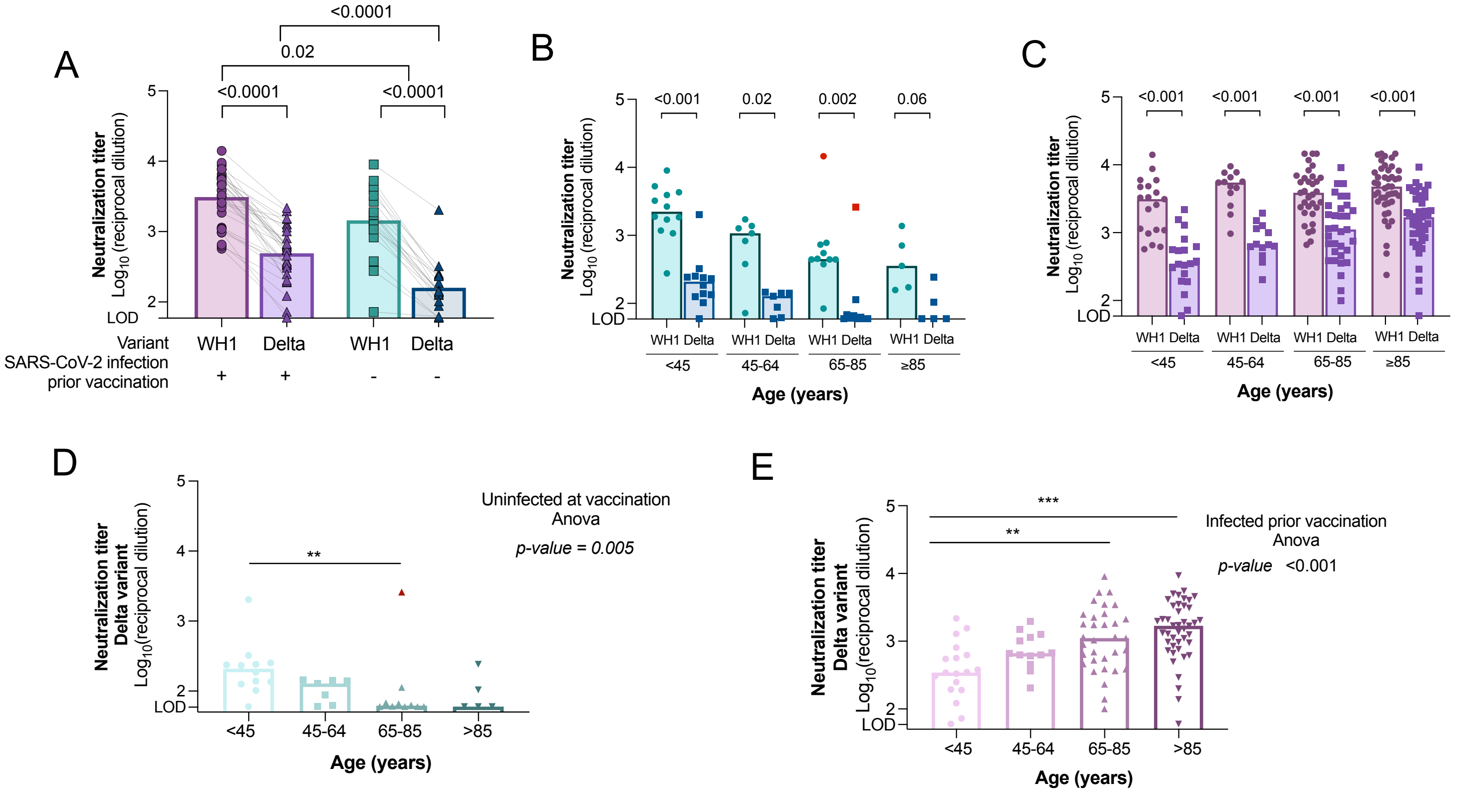
